# Frequency and profile of objective cognitive deficits in hospitalized patients recovering from COVID-19

**DOI:** 10.1101/2020.10.28.20221887

**Authors:** Abhishek Jaywant, W. Michael Vanderlind, George S. Alexopoulos, Chaya B. Fridman, Roy H. Perlis, Faith M. Gunning

## Abstract

**Background:** Cognitive impairment is common following critical illness. A number of case reports and case series have suggested that cognitive deficits occur in patients with COVID-19. This study evaluated the frequency, severity, and profile of cognitive dysfunction in hospitalized patients recovering from COVID-19.

**Methods:** We obtained and analyzed cross-sectional neuropsychological data from a cohort of N=57 patients participating in inpatient rehabilitation. Our primary outcome measure was the Brief Memory and Executive Test (BMET). We calculated the frequency of impairment based on clinician diagnosis and by the BMET subtests using age-normed classification of impairment. We explored associations with intubation and extubation as markers of illness severity and complications, as well as psychiatric diagnosis.

**Outcomes:** Our sample was 75% male, 61% non-white, with a mean age of 64.5 (SD = 13.9) years. Patients were evaluated at a mean of 43.2 days post-admission. 88% had documented hypoxemic respiratory failure and 77% required intubation. 81% of patients had cognitive impairment, ranging from mild to severe. Deficits were most common in working memory (55% of patients impaired), set-shifting (47%), divided attention (46%), and processing speed (40%). Executive dysfunction was not significantly associated with intubation length or the time from extubation to assessment, nor was it associated with the presence of a psychiatric diagnosis.

**Interpretation:** Medically stable inpatients recovering from COVID-19 commonly have deficits in attention and executive functions. These deficits were not significantly correlated with length of intubation or time since extubation. Findings provide an early benchmark for studying the evolution of cognitive difficulties after COVID-19 and suggest that easy to disseminate interventions that remediate attention and executive dysfunctions may be important in this population.

**Funding:** The authors have no funding for this study to report.

## Background

Cognitive deficits are frequent, persistent, and disabling following critical illness.^1,2^ They are increasingly recognized as a common complication of COVID-19. Multiple factors associated with the illness and its treatment may contribute to cognitive sequelae. These include hypoxia, ventilation, sedation, delirium, cerebrovascular events, and inflammation.^3–6^ To date, however, reports of cognitive functioning are largely limited to case reports and case series. Few investigations have used objective neuropsychological measures to quantify cognitive deficits,^7,8^ or to characterize the extent and profile of cognitive dysfunction during recovery from COVID-19. The time-sensitive need to understand and address cognitive deficits early in the disease course is underscored by the prevalence of COVID-19 coupled with the long-term cognitive and psychiatric complications that were associated with the coronaviruses that caused the first Severe Acute Respiratory Syndrome (SARS) and Middle East Respiratory Syndrome (MERS).^9^

In this study, we collected and analyzed data from neuropsychological assessments conducted in a cohort of patients recovering from COVID-19 on inpatient rehabilitation units at a large, tertiary care academic medical center in New York City. The goal of our study was to evaluate the frequency, severity, and profile of cognitive deficits in hospitalized and recovering patients with COVID-19. We investigated relationships between cognitive functioning and mechanical ventilation, known to be linked to long-term deficits following critical illness. We additionally explored the occurrence of psychiatric diagnoses and their association with cognitive functioning in COVID-19.

## Methods

### Sample

From April-July 2020, we administered neuropsychological measures to a cross-sectional cohort of N=57 hospitalized patients with COVID-19 participating in inpatient rehabilitation. All patients were initially admitted to the hospital for signs and symptoms of COVID-19 and were confirmed positive for SARS-CoV-2 via polymerase chain reaction (PCR). All patients were transferred to inpatient rehabilitation from intensive care, an acute inpatient medical/surgical floor, or an ICU stepdown unit. Rehabilitation occurred on a 22-bed general inpatient rehabilitation unit with 10 overflow beds added to meet demand and on a newly created 30-bed COVID Recovery Unit.^10^ Admission to the general rehabilitation unit was based on standard criteria, i.e. medically stable and able to tolerate three hours of rehabilitation therapy with reasonable expectation of functional gain. Patients admitted to the COVID Recovery Unit had to be medically stable, able to tolerate >30 minutes of therapy daily, and had an anticipated discharge home or to an acute or subacute rehabilitation facility; active delirium was an exclusion criterion. At the time of the neuropsychological assessment, all patients were medically stable and undergoing daily physical therapy, occupational therapy, and speech therapy as needed. Data reported here were extracted from neuropsychological reports via chart review after approval from the Weill Cornell Medicine Institutional Review Board.

### Measures

#### Demographic and Clinical Characteristics

We extracted from physician and neuropsychology notes in the medical record age, gender, race/ethnicity, length of hospitalization in days until the neuropsychological evaluation, intubation length in days, time since extubation until the neuropsychological assessment, and presence or absence of documented delirium in the ICU. To characterize functional disability in our sample, we extracted scores from the Activity Measure for Post-Acute Care (AM-PAC Inpatient Short Form)^11^ closest to admission to rehabilitation. The AM-PAC is a standard-of-care instrument administered by physical and occupational therapists to assess limitations in basic activities of daily living and basic mobility.

#### Cognitive Assessment

Patients were assessed at bedside by a clinical neuropsychologist or a neuropsychology postdoctoral fellow. Assessments occurred a mean of 6.6 days after patients’ transfer to the rehabilitation unit. Amongst cognitive domains, assessments focused on attention, executive functioning, and memory given the documented relation between these cognitive domains and functional outcome^12^ and because of the relationship between these domains and inflammation, vascular processes, hypoxia, and mood and anxiety symptoms.^13–16^

We report data from our core neuropsychological measure, the Brief Memory and Executive Test (BMET). The BMET is comprised of multiple subtests assessing aspects of executive functioning and memory that has demonstrated strong psychometric properties and sensitivity to impairment.^17^ The BMET has eight subtests that assess orientation, five-word immediate recall (working memory), five-word recall (delayed memory), five-word recognition (delayed recognition), rapid letter-number matching (divided attention), motor speed, rapid letter sequencing (visual attention and processing speed), and letter-number switching (set-shifting). The BMET letter-number switching task was substituted with the Oral Trail Making Test-B^18,19^ for patients who did not have adequate motor function, or for the Color Trails Test a relatively culture-fair version of trail making for non-English speakers.^20^ We used phone interpreters to conduct evaluations for 11 patients who did not speak English.

Our sample included patients who received at least one of these eight subtests. As some portions of the evaluations were unable to be completed in all patients due to ongoing medical care/rehabilitation, and due to patient motor limitations, the number of patients completing each subtest of the BMET varied. Patients who were not administered at least one of the above-mentioned measures were excluded from analysis; these measures were not administered when referral question was specific to psychiatric symptoms and treatment, patients preferred to focus on psychiatric symptoms or treatment, the neuropsychologist elected to limit evaluation to psychiatric symptoms, or the severity of cognitive deficits precluded administration of the full assessment.

#### Psychiatric Diagnosis

All patients were evaluated for depression, anxiety, and adjustment to disability by clinical interview. We extracted from the medical record the psychiatric diagnosis assigned by the neuropsychologist in the summary/impressions section of the report and related this to the presence of cognitive impairment.

### Statistical Analysis

Demographic and clinical characteristics were evaluated using descriptive statistics. To understand generalizability, we compared demographic and clinical characteristics of our analyzed sample with patients who were evaluated by neuropsychology but did not receive the BMET. We calculated the frequency of performance in the normal range, mild/borderline impaired range, and impaired range based on the normative sample and age-adjusted established cutoffs of the BMET (and Oral Trail Making Test and Color Trails Test when substitutions occurred). Impairment was classified separately for each BMET subtask. Mild/borderline was classified as a score <1 standard deviation below the age-adjusted normative mean, and impaired was classified as a score <2 standard deviation below the age-adjusted normative mean. We used impairment categories because of the absence of age-adjusted continuous Z-scores in the BMET normative sample. We used Pearson correlations to explore the association between performance on the letter-number matching (divided attention) subtest of the BMET and intubation length and time from extubation to the assessment as markers of illness severity. We chose to focus on the letter-number matching subtest because of the sensitivity of similar coding tests to executive dysfunction and functional disability.^21^ We used chi-square (χ^2^) tests to examine the association between cognitive impairment and psychiatric diagnosis.

## Results

### Sample demographic and clinical characteristics

Approximately 75% of the sample was male and 61% was non-white (Table 1). Mean age was 64·5 (SD = 13·9) years. 88% of patients had documented hypoxia/hypoxemic respiratory failure and 77% were treated with intubation and mechanical ventilation. 29% of patients were weaned off ventilation using tracheostomy. At the time of admission to rehabilitation, all patients were significantly limited (mean T-scores greater than 1.5 standard deviations below normative function) in basic mobility and activities of daily living as assessed by the AM-PAC. Patients who were evaluated with the BMET did not differ from those who were not administered the BMET (N=29) in age, length of hospitalization to assessment, intubation length, time from extubation to assessment, basic mobility, or activities of daily living (all *t*’s < 1·59, all *p*’s > ·11). Included and excluded patients did not differ in proportion of gender or race/ethnicity (all χ^2^ < ·98, all p’s > ·33).

**Table 1:** Demographics and Clinical Characteristics. Values represent mean (standard deviation) for continuous measures and N (%) for categorical measures

|  | Mean | SD |
| --- | --- | --- |
| **Age** | 64·5 | 13·9 |
| **Gender** |  |  |
| Male | 43 (75%) |  |
| Female | 14 (25%) |  |
| **Race/Ethnicity** |  |  |
| White | 22 (39%) |  |
| Latino/Hispanic | 16 (28%) |  |
| Black | 7 (12%) |  |
| Asian | 11 (19%) |  |
| Other | 1 (2%) |  |
| **Language of Assessment** |  |  |
| English | 46 (81%) |  |
| Spanish | 5 (9%) |  |
| Chinese (Mandarin, Cantonese, or regional dialect) | 4 (7%) |  |
| Other | 2 (4%) |  |
| **Documented Hypoxia/Hypoxemic Respiratory Failure** | 50 (88%) |  |
| **Intubated** | 44 (77%) |  |
| **Length of Intubation (Days)** | 13·2 | 10·1 |
| **Tracheostomy** | 16 (29%) |  |
| **Documented Delirium during Acute Hospitalization** | 37 (66%) |  |
| **AMPAC 6-clicks Basic Mobility (T-score)** | 34·4 | 8·57 |
| **AMPAC 6-clicks Daily Activities (T-score)** | 33·8 | 6·47 |
| **Time from Admission to Assessment (Days)** | 43·2 | 19·2 |
| **Time from Extubation to Assessment (Days)** | 26·8 | 14·0 |
| **Time from admission to Rehabilitation to Assessment (Days)** | 6·6 | 2·6 |
| **Cognitive Diagnosis Assigned by Clinician** |  |  |
| Normal cognitive functioning | 11 (19%) |  |
| Mild cognitive deficits | 27 (47%) |  |
| Moderate cognitive deficits | 14 (25%) |  |
| Severe cognitive deficits | 5 (9%) |  |
| **Psychiatric Diagnosis Assigned by Clinician** |  |  |
| No diagnosis (emotional function judged to be normative) | 34 (60%) |  |
| Adjustment disorder | 13 (23%) |  |
| Major Depressive Disorder | 2 (3%) |  |
| Unspecified anxiety or mood disorder | 6 (11%) |  |
| Pre-existing psychiatric illness still present | 2 (3%) |  |

**Note.** Cognitive diagnosis assigned by clinician was based on the BMET, adjunct neuropsychological measures, and clinical observations

### Cognitive functioning

The majority of patients (81%) were assessed by neuropsychologists as having at least some degree of cognitive deficits (Table 1). Mild cognitive impairment was most common, though moderate and severe cognitive impairment was also apparent in some patients. Figure 1 displays the percentage of patients classified as cognitively normal, mild/borderline, and impaired by subtest of the BMET, and shows that deficits were most commonly observed on subtests assessing attention and executive functions. For example, 46% of the sample exhibited impairment on the divided attention subtest. While rates of impairment on a simple motor speed task were low, impairment increased when a rapid visual attention and information processing speed component was added. Impairment increased further when a rapid set-shifting component was added.Immediate recall, which involves working memory, had greater percentage of impairment than did delayed recall or delayed recognition.

**Figure 1.**
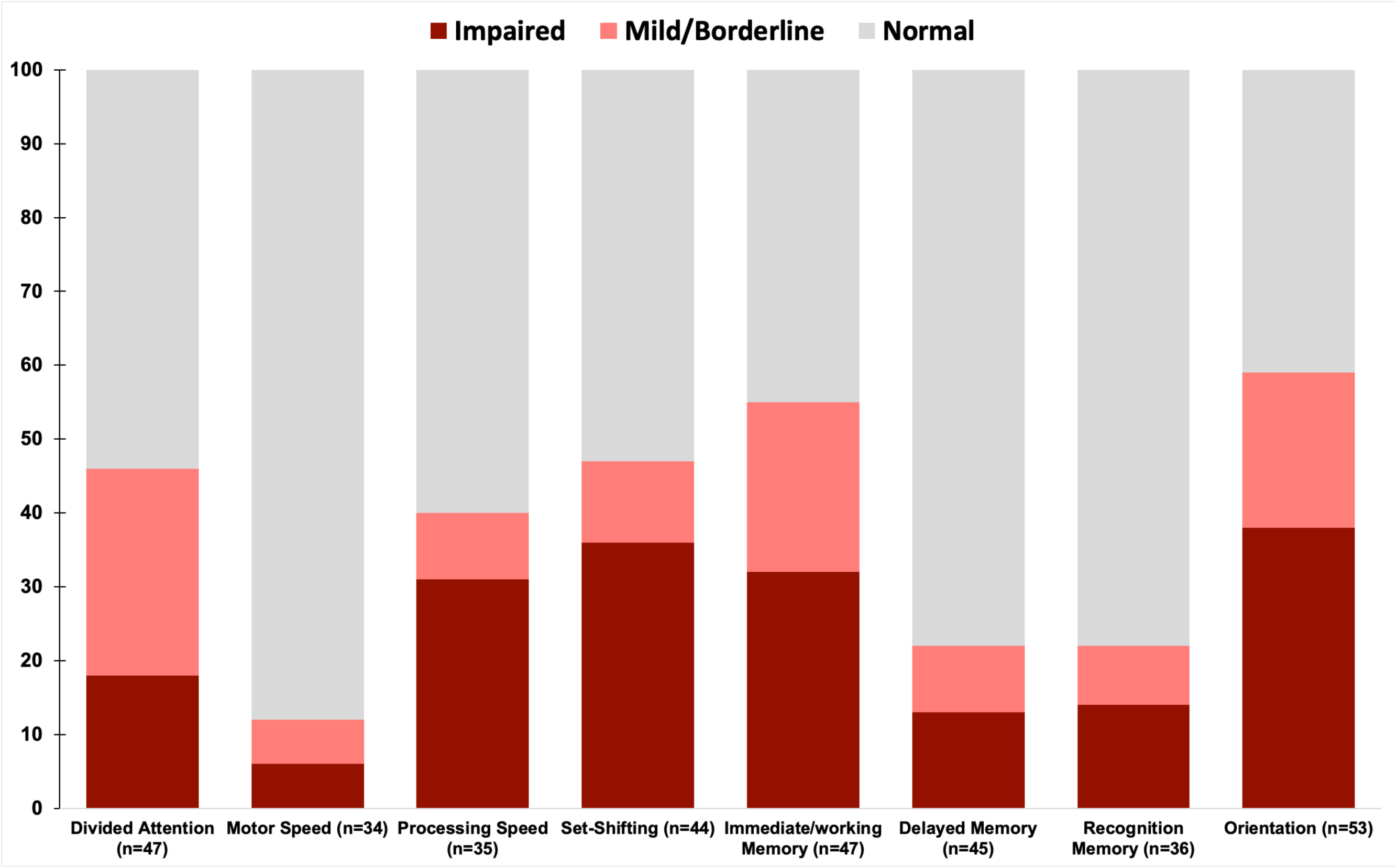
Percentage impairment by subtest of the Brief Memory and Executive Test. Classification was based on published norms for the BMET, with Mild/Borderline defined as <1 standard deviation below the age-adjusted norms and Impaired performance defined as <2 standard deviation below age-adjusted norms.

### Association between divided attention and intubation/extubation

We calculated Z-scores for the letter-number matching test relative to the overall BMET normative sample mean and standard deviation. Because the normative sample is not separated by age, we regressed the Z-score onto age and then used the unstandardized residuals in our analysis. As shown in Figure 2, divided attention was not significantly associated with length of intubation (*r* = -·12, *p*= ·49) or the time between extubation and assessment (*r* = -·25, *p* = ·19) after adjusting for age.

**Figure 2.**
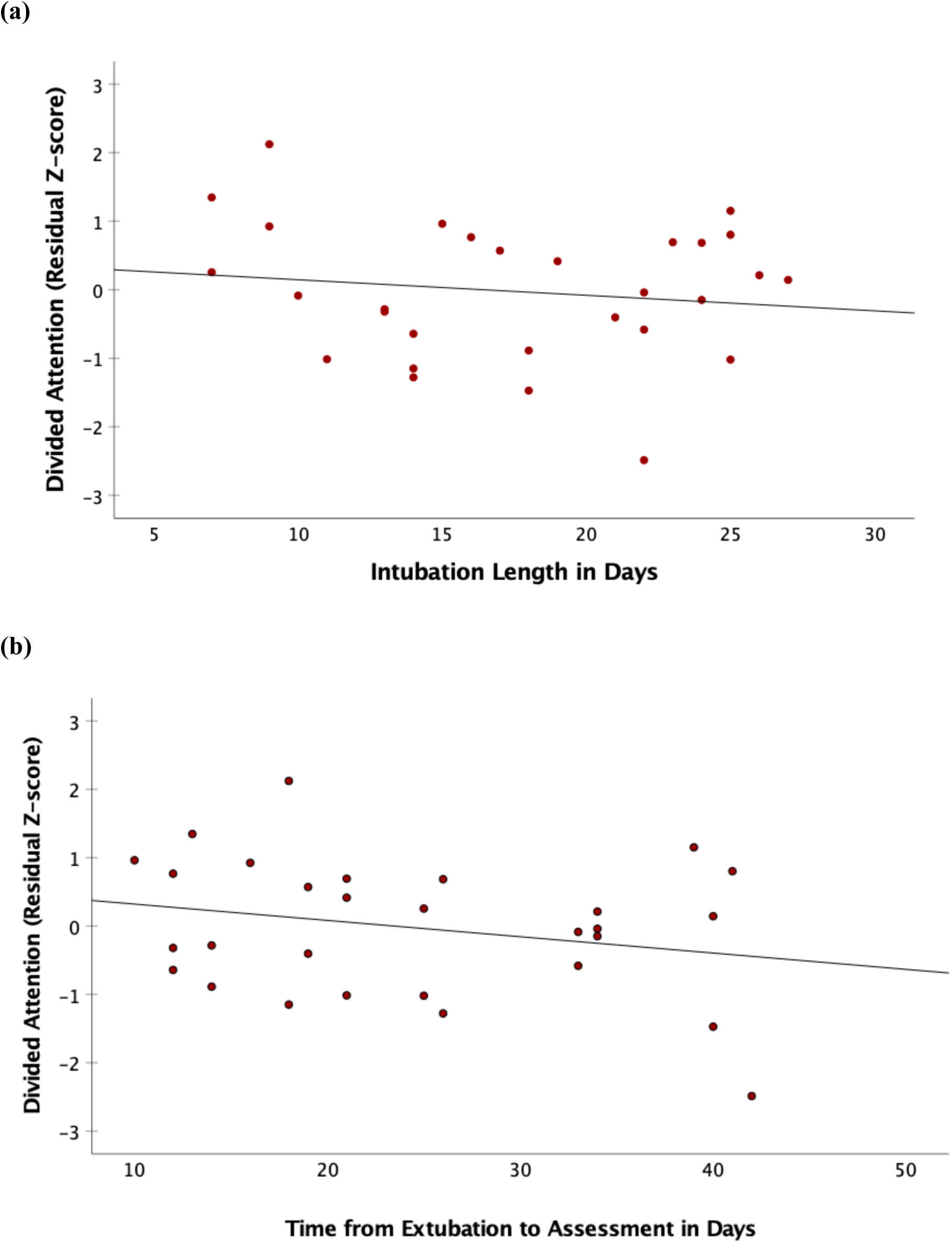
Association between Divided Attention and (a) Intubation Length in days and (b) Time Between Extubation and Assessment. Divided Attention (y-axis) is plotted as the residual Z-score relative to the normative sample after regressing out age.

### Association between cognition and psychiatric diagnosis

Table 1 also displays the frequency (percentage) of psychiatric diagnoses assigned by neuropsychologists. Overall, 58% of patients did not meet criteria for a psychiatric diagnosis The remaining patients were found to have adjustment disorders with anxiety, depression, or mixed anxiety and depression. Some were assigned an unspecified mood/anxiety disorder, while a small number met clinical criteria for major depressive disorder. Using a 2×2 χ^2^ test, there was no significant relationship between cognition (normal, impaired) and psychiatric symptoms (normal, symptomatic), χ^2^ = ·92, *df* =1, *p* = ·34.

## Discussion

The main finding of this study is that 81% of medically stable and recovering COVID-19 inpatients had objectively-documented cognitive deficits that ranged from mild to severe. Attention and executive functions were most affected. That is, rates of impairment increased as tasks placed greater demands on executive functions. Divided attention, set-shifting, and processing speed had relatively high rates of impairment. Similarly, immediate recall—which places high demands on working memory—had a high rate of impairment, whereas delayed memory and recognition memory were infrequently impaired. To our knowledge this is the first report of objectively measured cognitive symptom profiles in a well-characterized sample of individuals recovering from COVID-19.

Our findings extend those of a prior study of 29 community-dwelling adults in China recovered from COVID-19 that found a deficit in sustained attention,^8^ though in that study the disease and treatment characteristics were not described. The early evidence in COVID-19 thus indicates that deficits in attention and executive functions are more common than deficits in memory. Our findings suggest the involvement of brain regions relevant to executive control processes such as the prefrontal cortex, parietal cortex, cingulate cortex, and striatum.

We explored the association of a sensitive measure of executive function, divided attention, with intubation duration and time between extubation and assessment. After controlling for age, divided attention was not associated with either intubation duration or time from extubation. Although longer duration of mechanical ventilation is known to predict worse long-term functional outcome after critical illness,^22^ we did not detect a significant relationship with executive dysfunction in this cohort. Intubation durations after COVID-19 have been significantly longer than in prior acute respiratory distress syndromes and it is thus possible that after a certain threshold passed by COVID-19 patients, cognitive deficits may occur irrespective of intubation duration. The absence of a positive association between time from extubation to assessment and divided attention was somewhat surprising given that one would expect better cognitive performance further from the removal of mechanical ventilation. The lack of association between time from extubation and divided attention suggests our findings may not simply be an artifact of resolving acute alterations in mental status.

Whether the frequency and severity of cognitive deficits in COVID-19 is unique from the known cognitive dysfunction that occurs more generally following critical illness, ICU admission, and respiratory distress is yet to be determined. In (non-COVID-19) critically ill patients, delirium in the hospital occurs at a frequency of >70% of patients.^1^ At discharge, the prevalence of cognitive dysfunction is approximately 80% for survivors of acute respiratory distress syndrome,^23^ which is similar to our sample in which 81% exhibited cognitive impairment. At three months, 40% of critically ill patients have at least mild cognitive deficits and 26% have moderate cognitive deficits.^2^ Assessed at an earlier time point—on average 43 days after admission—we found similar percentages of COVID-19 patients with mild and moderate impairment. Our finding of predominant executive functioning deficits is also consistent with known executive dysfunction after critical illness.^24^ Although there are similarities to the existing literature on critical illness, there are factors unique to COVID-19 that may increase the potential for long-term cognitive dysfunction and disability. Patients have been intubated and ventilated for long durations and cerebrovascular complications have been shown to be common.^25^ The long-term evolution of cognitive deficits after COVID-19 and their unique characteristics are important questions for continued research because of impact of cognitive dysfunction on long-term functional disability.^26^

We found that 60% of our sample was assessed as having absent or normal levels of anxiety and depressed mood (i.e., were not assigned a psychiatric diagnosis). Patients who were symptomatic primarily exhibited adjustment-related anxiety and depressed mood while only three patients met criteria for major depressive disorder. In an exploratory analysis, cognitive impairment was not associated with psychiatric diagnosis; however, the lack of association may be affected in part by the neuropsychologists’ decision not to administer the BMET to patients for whom assessing and treating psychiatric symptoms was the focus of the evaluation. After critical illness, executive functioning deficits increase risk for future depressive symptoms.^27^ Thus, it will be important to examine if patients with executive dysfunction develop depressive syndromes after hospital discharge.

The generalizability of the current study is limited by the evaluation of a subset of patients undergoing rehabilitation in a single hospital; however, those tested with the BMET did not differ from those patients evaluated by neuropsychology who were not administered the BMET, and the demographics of our sample was consistent with known gender, racial, and ethnic differences in COVID-19 infection.^28,29^ Although we were not able to administer a comprehensive neuropsychological evaluation in the inpatient setting, the BMET enabled the quantification of the severity and pattern of deficits in aspects of attention, executive functions, and memory. That different subtests were given at different frequencies reflects the reality of neuropsychological evaluation in a busy inpatient setting at the height of the initial surge of COVID-19 cases in New York City. Despite limitations, our study is among the largest to date using objective and standardized neuropsychological assessment. Because our neuropsychologists were embedded on the rehabilitation units, we were able to administer in-person cognitive assessments while following designated infection control procedure. Our results may help to establish a benchmark for early cognitive dysfunction after severe COVID-19 illness.

## Conclusion

In summary, we show that medically stable inpatients recovering from COVID-19 commonly have impairments in attention and executive functions (i.e., working memory, divided attention, set-shifting). Divided attention was not associated with intubation duration or time from extubation to assessment. While intubation duration may be one measure of a more severe course, other markers of illness severity may be associated with cognitive sequelae. Our results provide an early benchmark for studying the evolution of cognitive difficulties in recovering COVID-19 patients. They also highlight the importance of studying interventions that target attention and executive functioning after COVID-19. Given the prevalence of COVID-19, targeting these deficits through scalable cognitive interventions that have been demonstrated to improve similar deficits and can be widely disseminated in patients’ homes through reliance on technology^30^ may support optimal cognitive and functional outcomes.

## Data Availability

Data for this study are not currently available.

## Acknowledgements

The authors did not receive any funding to complete this study. We thank Ruchi Patel, MA OTR/L, Gargi Doulatani, BA, and Karen Wen, BA, for their assistance in data extraction and management. We are grateful for the tireless physicians, nurses, and rehabilitation therapists who worked with us to assist COVID-19 survivors in their rehabilitation and recovery.

## Author contributions

A.J. conducted clinical interviews and neuropsychological evaluations, extracted and verified the data, conducted data analyses, and wrote the manuscript. W.M.V. conducted clinical interviews and neuropsychological evaluations, extracted and verified the data, and edited the manuscript. G.S.A. and C.B.F. provided input in measure selection, interpretation of results, and manuscript writing. R.H.P. and F.M.G. assisted in data analysis, interpretation of results, and manuscript writing.

